# Rapid fluoroquinolone resistance detection in *Pseudomonas aeruginosa* using mismatch amplification mutation assay-based real-time PCR

**DOI:** 10.1101/2021.12.12.21266792

**Authors:** Danielle E. Madden, Kate L. McCarthy, Scott C. Bell, Olusola Olagoke, Timothy Baird, Jane Neill, Kay A. Ramsay, Timothy J. Kidd, Adam G. Stewart, Shradha Subedi, Keat Choong, Tamieka A. Fraser, Derek S. Sarovich, Erin P. Price

## Abstract

**Background:** Antimicrobial resistance (AMR) is an ever-increasing global health concern. One crucial facet in tackling the AMR epidemic is earlier and more accurate AMR diagnosis, particularly in the dangerous and highly multi-drug resistant ESKAPE pathogen, *Pseudomonas aeruginosa*.

**Objectives:** We aimed to develop two SYBR Green-based mismatch amplification mutation assays (SYBR-MAMAs) targeting GyrA T83I (*gyrA*248), and GyrA D87N, D87Y, and D87H (*gyrA*259). Together, these variants cause the majority of fluoroquinolone (FQ) AMR in *P. aeruginosa*.

**Methods:** Following assay validation, the *gyrA*248 and *gyrA*259 SYBR-MAMAs were tested on 84 clinical *P. aeruginosa* isolates from Queensland, Australia, 45 of which demonstrated intermediate/full ciprofloxacin resistance according to antimicrobial susceptibility testing.

**Results:** Our two SYBR-MAMAs correctly predicted an AMR phenotype in the majority (84%) of isolates with intermediate/full FQ resistance. Importantly, all FQ-sensitive strains were predicted to have a sensitive phenotype. Whole-genome sequencing confirmed 100% concordance with SYBR-MAMA genotypes.

**Conclusions:** Our GyrA SYBR-MAMAs provide a rapid and cost-effective method for same-day identification of FQ AMR in *P. aeruginosa*. An additional SYBR-MAMA targeting the GyrB S466Y/S466F variants would increase FQ AMR prediction to 91%. Clinical implementation of our assays will permit more timely treatment alterations in cases where decreased FQ susceptibility is identified, leading to improved patient outcomes and antimicrobial stewardship.

## Background

The ESKAPE pathogen, *P. aeruginosa*, has a remarkable capacity to develop AMR towards all clinically-relevant antibiotic classes (1). This bacterium can cause life-threatening infections, particularly in people with wounds, cancer, or chronic respiratory diseases such as cystic fibrosis (CF) or chronic obstructive pulmonary disease (COPD) (2).

Rapid, affordable, accessible, and accurate AMR diagnosis is crucial in the battle against ESKAPE pathogens (1). However, few diagnostic tests exist (3) for rapidly and inexpensively characterising AMR-conferring single-nucleotide polymorphisms (SNPs) in *P. aeruginosa*, a striking knowledge gap given that SNPs confer AMR towards anti-pseudomonal drugs such as fluoroquinolones (FQs) (4).

FQs (predominantly ciprofloxacin [CIP]) have proven clinically useful for treating *P. aeruginosa* infections (5). Yet, upon exposure, *P. aeruginosa* often develops FQ resistance (FQr). Codon-altering mutations within the GyrA quinolone-resistance-determining region (QRDR) can confer an intermediate (CIPi) or fully resistant (CIPr) CIP phenotype (6, 7); in contrast, *gyrB, parC, parE* and *nfxB* typically require ≥2 mutations to impart CIPi/CIPr (8).

Due to the single-step nature of QRDR mutations in conferring CIPi/CIPr, and their prevalence in clinical isolates (9), we targeted the two most common QRDR SNPs, *gyrA*248 (4) and *gyrA*259 (10), for assay development. We chose SYBR Green-based mismatch amplification mutation assay (SYBR-MAMA), an inexpensive (∼AUD$1-2/assay when run in duplicate), simple, rapid (∼1h turnaround time) and scalable method (11-15) that exploits the differential efficiency of allele-specific amplification for SNP interrogation; this efficiency disparity can be observed in real-time by measuring the difference in cycles-to-threshold (ΔC_T_) (12, 14, 15).

## Methods

Eighty-four *P. aeruginosa* isolates from Southeast Queensland, Australia, were examined: 42 from sputum derived from adults with CF and chronic *P. aeruginosa* infection admitted to The Prince Charles Hospital between 2017 and 2019 (16); 35 bloodstream isolates retrieved from adults admitted to several public and private hospitals in Brisbane between 2008 and 2011 (17), three from COPD sputum, collected during in-home community nurse visits in the Sunshine Coast region (16); one from an adult with non-CF bronchiectasis collected in 2017, and one from an adult with urinary tract infection collected in 2018, both during admission to the Sunshine Coast University Hospital (Table 1). One ulcer and one ear infection isolate, both from Brisbane, were obtained from the 1,000 International *P. aeruginosa* Consortium collection (18). Ethics approvals were obtained as previously described (16, 17, 19).

**Table 1.**
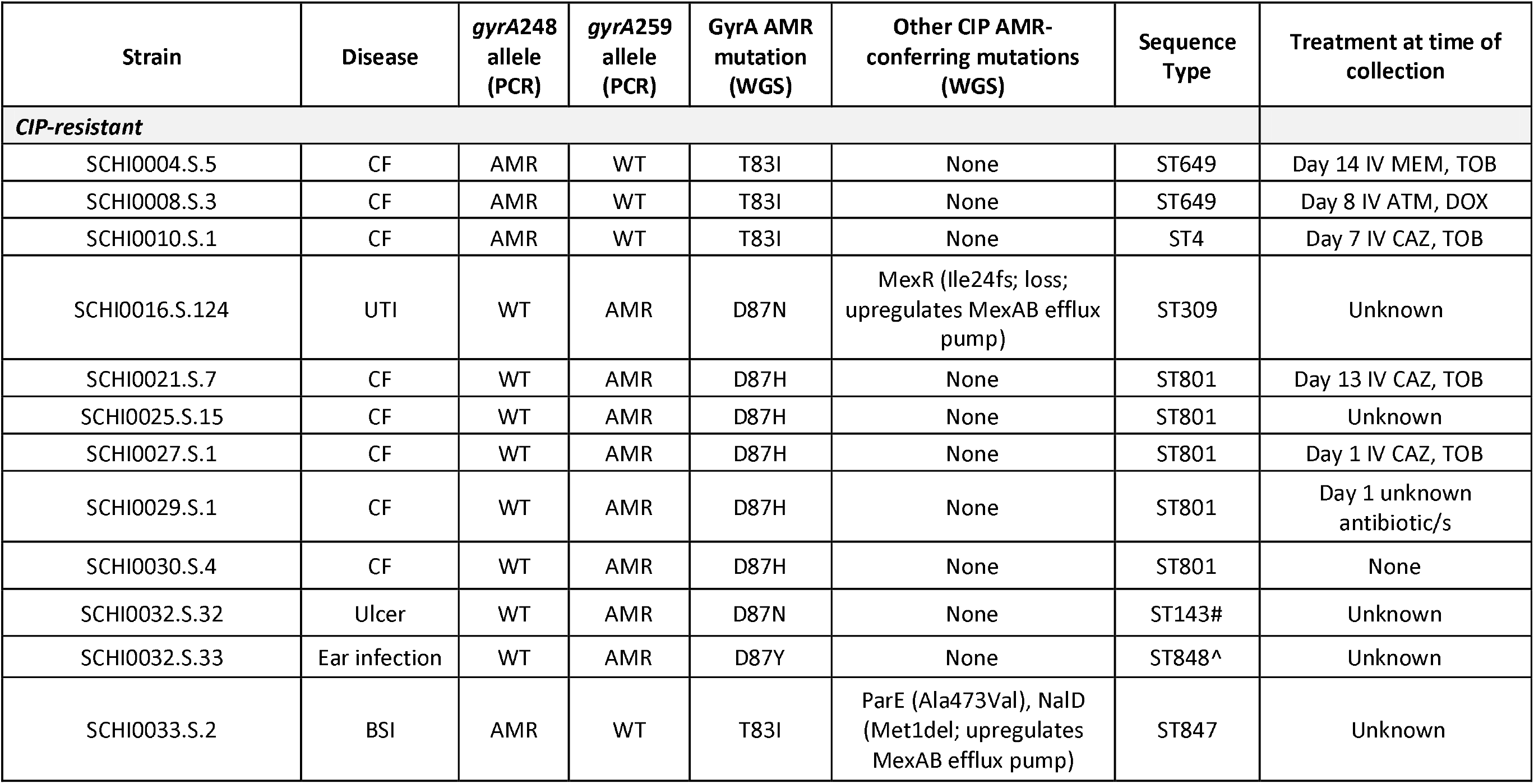
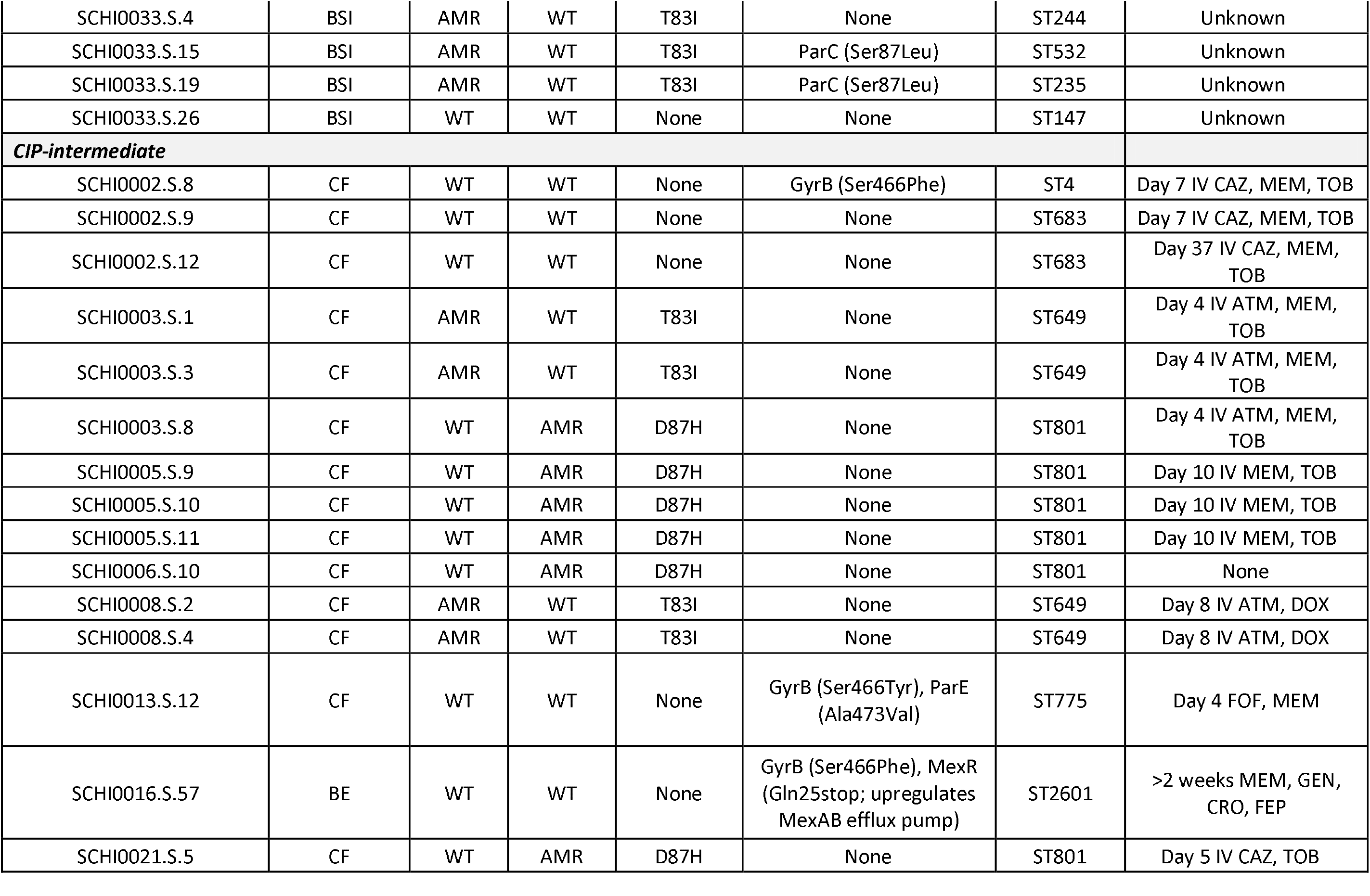
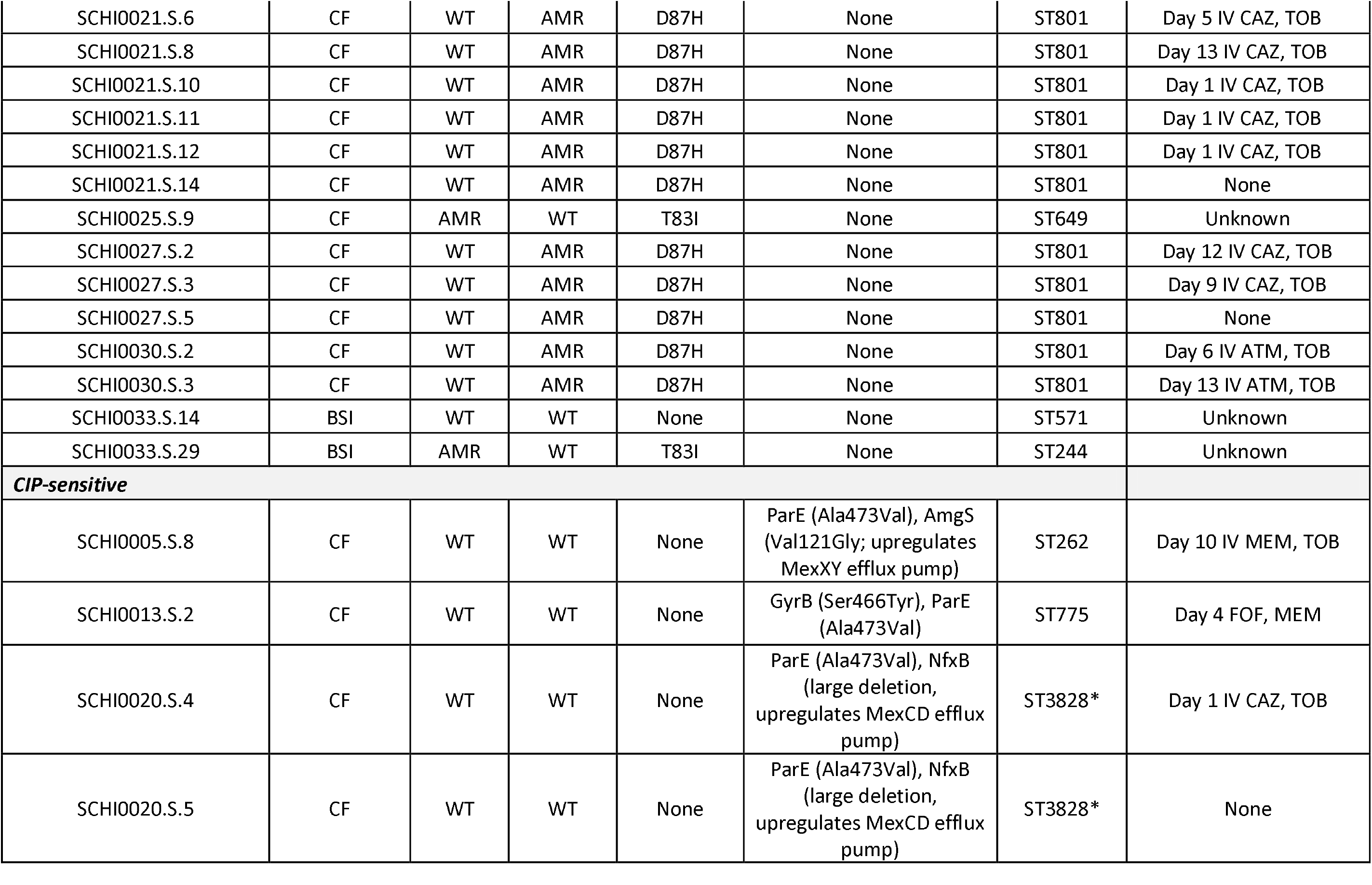
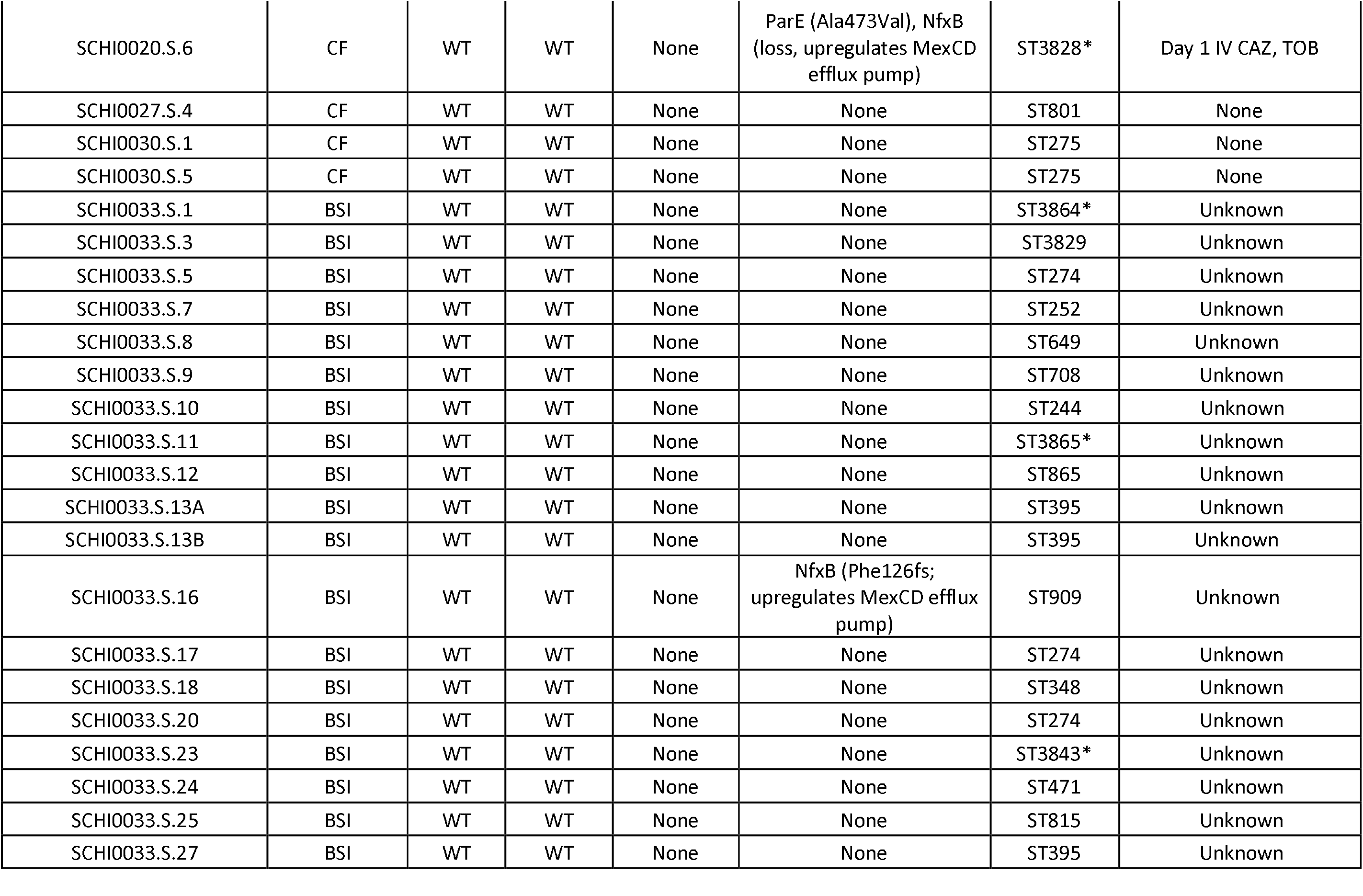
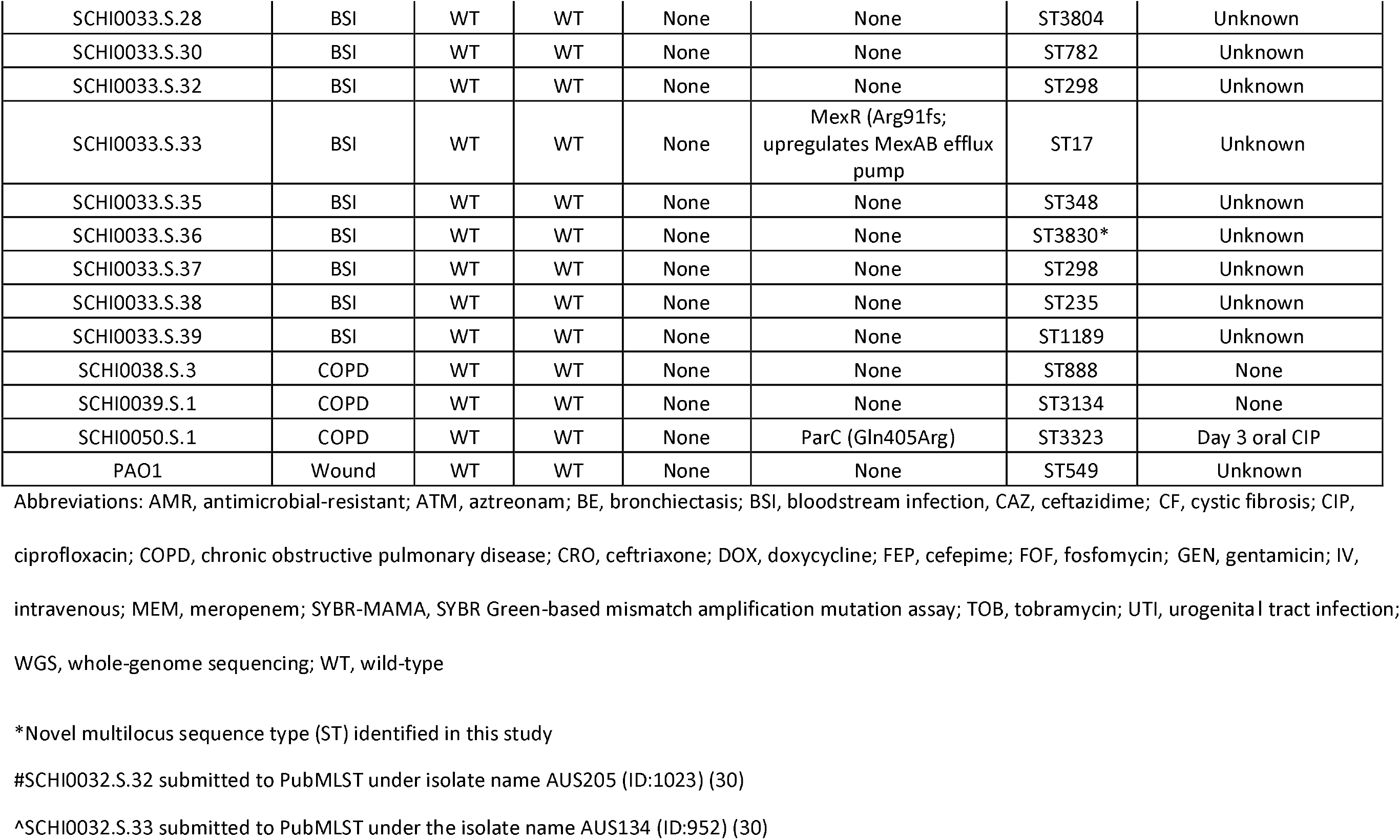
*Pseudomonas aeruginosa* isolates used in this study, and their DNA gyrase A (GyrA) SYBR-MAMA PCR genotypes, whole-genome sequencing (WGS) results, multilocus sequence types, and treatment details.

Strains were isolated using MacConkey agar (Oxoid, VIC, Australia) incubated at 37⍰C for 24h and confirmed as *P. aeruginosa* by *ecfX* real-time PCR (20). Susceptibility towards CIP [5μg], levofloxacin (LEV; 5 μg), moxifloxacin (MFX; 5 μg), and ofloxacin (OFX; 5 μg) was determined by disc diffusion (Edwards Group, QLD, Australia) using CLSI M100S-Ed27:2017 guidelines (Table 2).

**Table 2.**
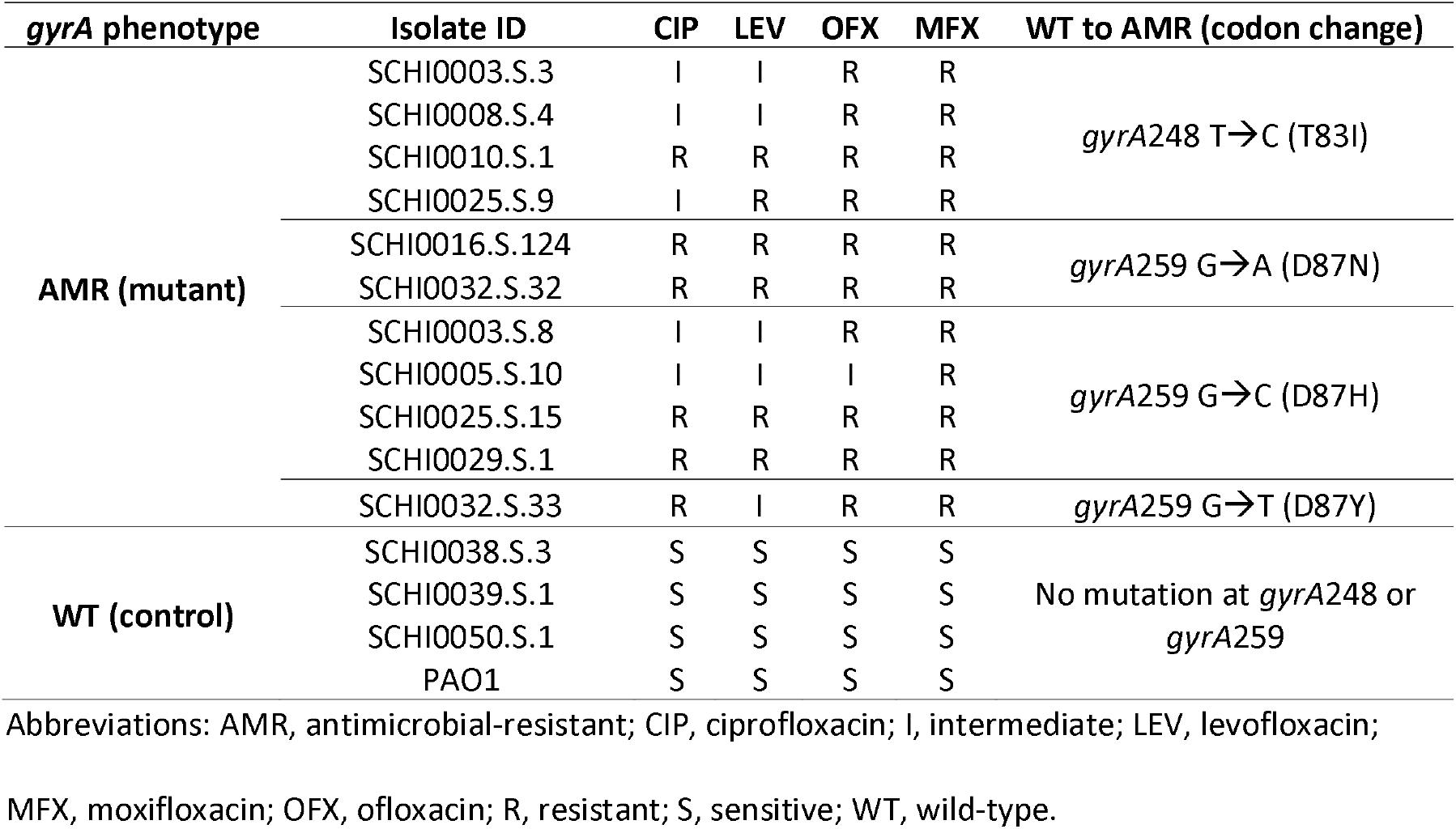
*Pseudomonas aeruginosa* isolates used for *gyrA*248 and *gyrA*259 SYBR-MAMA assay validation, and associated fluoroquinolone-class antibiotic disc diffusion data.

Isolates were DNA-extracted using the DNeasy kit (Qiagen, Chadstone Centre, VIC, Australia), followed by Illumina paired-end whole-genome sequencing (WGS) (19). Quality-filtered reads (21) were assembled with MGAP v1.1 (https://github.com/dsarov/MGAP---Microbial-Genome-Assembler-Pipeline) or SPAdes (22). AMR prediction was undertaken using a *P. aeruginosa*-specific ARDaP (23) database. Assemblies were deposited into the PubMLST database (https://pubmlst.org/organisms/pseudomonas-aeruginosa). Illumina data are available via BioProject PRJNA761496. We also tested rapid DNA extraction of 10 representative strains using the 5% chelex-100 rapid heat soak method (24), followed by a 1:10 dilution in molecular-grade H_2_O prior to PCR.

SYBR-MAMA primers were assessed *in silico* for dimer formation and specificity as described previously (25). For the *gyrA*248 SYBR-MAMA, gyrA248_T_AMR amplifies the CIPi/CIPr-conferring T83I allele, whereas gyrA248_C_WT amplifies the wild-type allele (Table 3). For the *gyrA*259 SYBR-MAMA, gyrA259_D_AMR amplifies mutant alleles at position 259 (D87N, D87Y, D87H), all of which confer CIPi/CIPr (4, 8), whereas gyrA259_G_WT amplifies the wild-type allele (10). PCRs consisted of 1× SsoAdvanced Universal SYBR Green Supermix (Bio-Rad, NSW, Australia), 0.2μM primers, 1μL template, and PCR-grade H_2_O, to 5μL. Thermocycling comprised 95⍰C for 2min, followed by 40 cycles of 95⍰C for 5s and 60⍰C for 5sec. Strains used to validate *gyrA* 248 and 259 alleles are listed in Table 2.

**Table 3.**
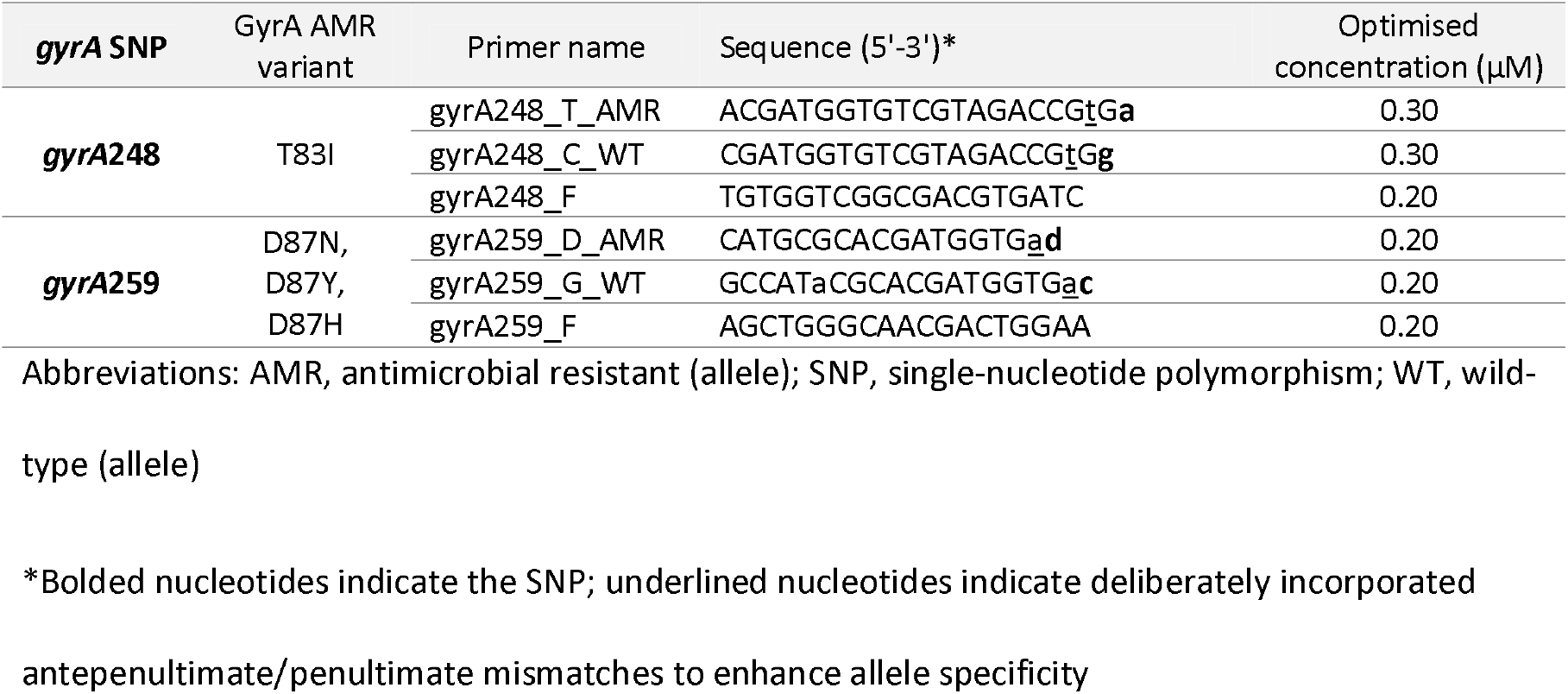
SYBR Green-based mismatch amplification mutation assay (SYBR-MAMA) primers designed in this study.

## Results

SYBR-MAMAs were screened across the 85 genome-sequenced *P. aeruginosa* isolates, comprising 40 CIP-sensitive (including PAO1), 29 CIPi, and 16 CIPr strains (Table 1). Of these, only one, SCHI0050.S.1, was derived from a participant receiving FQ (CIP) treatment, and this isolate was CIP-sensitive (Table 1). Unexpectedly, none of the 45 CIPi/CIPr strains were from participants known to be receiving contemporaneous FQ antibiotics (Table 1), although we were unable to investigate historical FQ exposure due to ethical limitations on participant data collection. It is therefore possible that some of our participants have previously received FQ antibiotics in the weeks or months prior to our sample collection. Alternatively, given that most of our participants were hospitalised, another possibility is that the CIPi/CIPr strains were nosocomially acquired, either from other admitted patients who had or were being treated with FQs, or from the hospital environment.

The *gyrA*248 SYBR-MAMA robustly discriminated GyrA T83I from wild-type strains, with matched alleles consistently amplifying earlier than mismatched counterparts (T83I ΔC_T_=4.0±0.03 vs. wild-type ΔC_T_=7.6±0.1 [Figures 1A and 1B]). Four tested GyrA T83I strains also demonstrated intermediate or full resistance towards LEV, MFX, and OFX (Table 2), confirming the importance of this variant in broader FQr. T83I is considered the most common GyrA variant in CIPi/CIPr strains (4, 9, 26); for example, two Japanese studies reported 82% (60/73) (27) and 75% (112/150) (28) T83I prevalence among CIPi/CIPr isolates, and a Vietnamese study reported 54% (76/141) prevalence (9). In our dataset, T83I was detected in 29% (13/45) CIPi/CIPr strains and 0% (0/40) CIP-sensitive strains (Table 1), suggesting that T83I is an important, but not dominant, cause of CIPi/CIPr in Australian isolates, although testing across a broader isolate collection is required to confirm this observation.

**Figure 1.**
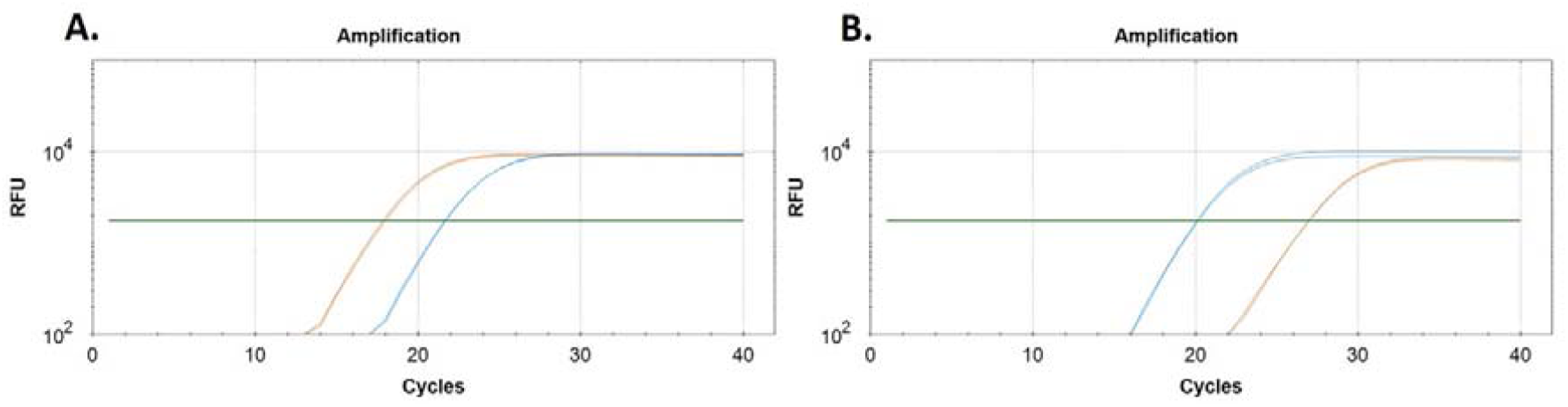
SYBR Green-based mismatch amplification mutation assay (SYBR-MAMA) interrogation of the *gyrA*248 (T→C) biallelic single-nucleotide polymorphism, resulting in a threonine to isoleucine substitution at position 83 (T83I) in certain fluoroquinolone (FQ)-resistant *P. aeruginosa* isolates. *gyrA*248 SYBR-MAMA performance in: **A**. SCHI0010.S.1 (FQ-resistant, encodes T83I); **B**. SCHI0038.S.3 (FQ-sensitive wild-type isolate). Orange, antimicrobial-resistant allele; blue, wild-type allele. No-template controls did not amplify. All samples were run in duplicate.

Like *gyrA*248, there was clear discrimination between AMR and wild-type genotypes for the *gyrA*259 SYBR-MAMA, with AMR alleles amplifying earlier in AMR-encoding strains (D87Y ΔC_T_=14.7 [Figure 2A]; D87N ΔC_T_=9.6±0.04 [Figure 2B]; D87H ΔC_T_=13.5±0.2 [Figure 2C]), and vice versa for wild-type strains (ΔC_T_=10.8; Figure 2D). *gyrA*259 AMR was detected in 56% (25/45) CIPi/CIPr and 0% (0/40) CIP-sensitive strains. The degenerate nature of the *gyrA*259 AMR primer has the advantage of enabling all four nucleotide variants to be detected using just two reactions; it does not require each variant to be tested individually. We therefore used WGS to differentiate each of the *gyrA*259 AMR variants to determine their prevalence. D87H was most common according to WGS, accounting for 88% (22/25) *gyrA*259 AMR strains (Table 1). In no instance did we observe a strain encoding both *gyrA*248 and *gyrA*259 AMR variants.

**Figure 2.**
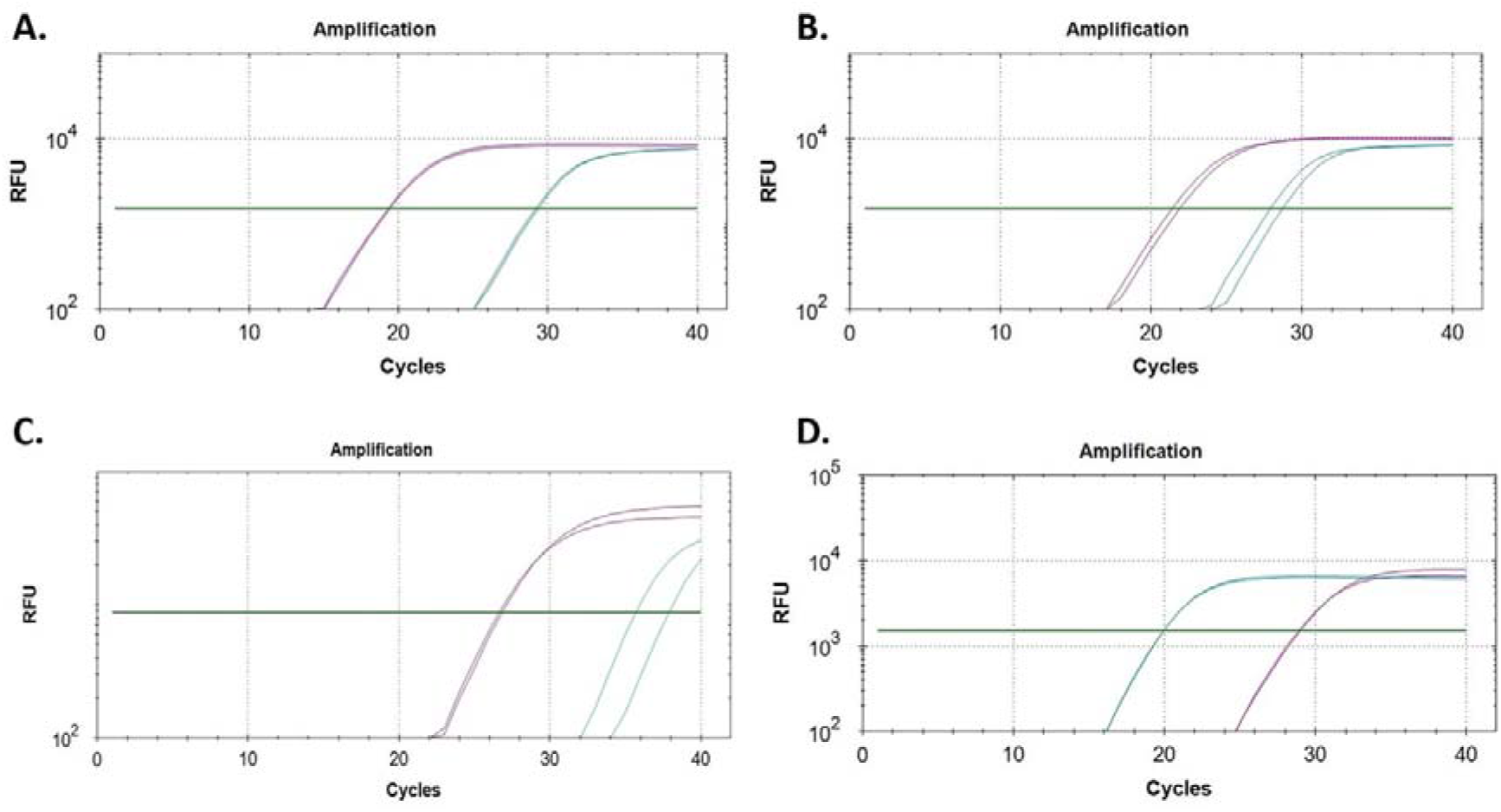
SYBR Green-based mismatch amplification mutation assay (SYBR-MAMA) interrogation of the *gyrA*259 (G→A/C/T) tetra-allelic single-nucleotide polymorphism, resulting in aspartate to asparagine (D87N), histidine (D87H), or tyrosine (D87Y) substitutions, respectively, at position 87 in certain fluoroquinolone (FQ)-resistant *Pseudomonas aeruginosa* isolates. *gyrA*259 SYBR-MAMA performance in: **A**. SCHI0005.S.10 (intermediate resistance towards FQs, encodes D87H); **B**. SCHI0032.S.32 (FQ-resistant, encodes D87N); **C**. SCHI0032.S.33 (FQ-resistant, encodes D87Y); and **D**. SCHI0038.S.3 (FQ-sensitive wild-type isolate). Purple, antimicrobial-resistant allele; aqua, wild-type allele. No-template controls did not amplify. All samples were run in duplicate.

Like T83I, seven tested strains encoding D87Y, D87N and D87H all exhibited intermediate or full LEV, MFX, and OFX resistance (Table 2). The *gyrA* mutations had a larger impact on MFX and OFX (causing AMR) compared with CIP and LEV (causing intermediate resistance) (Table 2). This observation supports the hypothesis that additional mutation/s are sometimes required to confer AMR towards CIP and LEV (10).

We tested our two SYBR-MAMAs against rapid chelex-extracted heat soak DNA. In all instances, isolates yielded excellent, early amplification, and genotyped as expected. Although not tested in this study, further time savings could be made by performing SYBR-MAMAs using colony PCR (29); however, in our experience, this approach requires a high degree of skill due to the inhibitory nature of total cellular extracts in PCR, and typically results in a proportion of PCR failures, even with skilled operators. This issue is particularly acute when using low (e.g. 5 μL) reaction volumes, as was used in our study to minimise costs. Therefore, we recommend that chelex extractions followed by 1:10 dilution be performed where rapid DNA extraction is desired to ensure 100% amplification success.

Of the seven CIPi/CIPr strains that did not encode *gyrA*248 or *gyrA*259 AMR variants, three had GyrB Ser466Phe or Ser466Tyr missense mutations with or without other FQr-conferring mutations (ParE Ala473Val, MexR Gln25Stop), whereas four had no known FQr determinants (Table 1). A possible explanation for these latter strains is phenotype misclassification as 3/4 strains sat on the CIPi/sensitivity breakpoint. Although not performed in this study, determining the minimum inhibitory concentration of these strains using Etests or broth microdilution may help resolve these borderline phenotypes.

## Conclusions

Our two *gyrA*248 and *gyrA*259 SYBR-MAMAs detected FQ non-susceptibility in 84% Australian CIPi/CIPr strains. Importantly, all FQ-sensitive strains yielded wild-type genotypes for both assays, demonstrating 100% specificity. Our two *gyrA* SYBR-MAMAs thus provide a same-day, inexpensive, simple, and accurate tool for detecting the two most prevalent causes of FQ non-susceptibility in *P. aeruginosa*. An additional SYBR-MAMA targeting GyrB Ser466Phe and Ser466Tyr would increase detection of FQ non-susceptibility to 91% without loss of specificity. Conversion of our assays to single-tube Melt-MAMA format (11) would further decrease assay costs and PCR setup time.

## Data Availability

All genome sequence data generated in this study are available from NCBI BioProject PRJNA761496.

## Acknowledgements

We thank Patrick Harris at UQ Centre for Clinical Research for helpful discussions.

## Transparency declaration

The authors declare no competing interests. The funders had no role in study design, data acquisition, analysis, interpretation, writing or submission of the manuscript.

